# Trends in GP dispensing of thickeners in the NHS England, 2019-24

**DOI:** 10.1101/2025.09.04.25334883

**Authors:** Arlene McCurtin, Lindsey Collins, Owen Doody, Dervla Kelly, Tracy Lazenby-Paterson, Dominika Lisiecka, Alison Smith, Shaun O’Keeffe

## Abstract

**Introduction:** Oropharyngeal dysphagia is common and often results in aspiration. Thickeners are widely used to thicken liquids to slow the liquid bolus and prevent aspiration and aspiration pneumonia, despite multiple concerns about their use. To date, there is little data on prescribing/dispensing practices. This study aims to provide the first overview of thickener dispensing practices, serve as foundational material for parties interested in research, practice and policy making, and guide further work in this area.

**Method:** We explored English general practitioner thickener dispensing practices for the five-year period 2019-2024 via the NHS Business Services Authority ePACT2 database and using descriptive statistics and linear regression to analyse the data. We hypothesized that with a large data set, it would be possible to detect trends in dispensing. for the

**Results:** Between 2019 and 2024, nearly three million thickening items were dispensed in primary care in England at a cost of nearly 90 million sterling. Thickened liquids is primarily an intervention for older people although there was a significant decreasing trend in dispensed thickener items in adult populations. Thickener dispensing has increased in the youngest age group (0-1-years-old). The cost of thickeners to the NHS has risen in the last year under study despite the significant decline in use observed for most age groups. Of dispensed items over the five-year period, the vast majority were gum-based.

**Conclusion:** While the data provided in this study is limited to GP dispensed thickeners in England, it provides researchers, clinicians and policy makers with initial information on dispensing practices. From the available data, a decreasing dispensing trend is seen in the context of rising costs, older people comprise the population who primarily receive thickeners and gum-based products dominate the market. This study raises a number of questions regarding the use of thickeners for people with swallowing problems.

## Introduction

Oropharyngeal dysphagia is common, with one in 17 adults affected {1}. The occurrence is particularly high in older people, specifically those in long-term residential care (LTRC), with a prevalence of 56% identified in a recent study {2}. Dysphagia is defined as a functional impairment that either prevents or limits the intake of food and fluids, and which makes swallowing unsafe, inefficient, uncomfortable or affects quality of life {3}. It increases the risk of penetration and/or aspiration of food and fluid into the airways. Health care professionals widely use thickeners to try and prevent aspiration by thickening liquids with a range of starches and gums in order to increase viscosity and slow the flow of the bolus. Thickeners are ideally recommended after a clinical swallow exam, which is performed by a registered and dysphagia trained Speech and Language Therapist (SLT), after which it is usually prescribed by the appropriate physician. While it may also be prescribed by nurse prescribers, the evidence for nurse prescribing of thickeners is currently limited {4}. SLTs have typically demonstrated high levels of use of thickeners {5,6}, however, a drift towards decreased recommendations is suggested by the results of a recent international survey, which highlights that only 20% of SLTs frequently recommend the intervention {7}.

The widespread use of thickeners for people with swallowing problems has come under increased scrutiny and criticism in recent years. Although thickeners are used with the ultimate goal of preventing aspiration pneumonia, there is little evidence that they achieve this {8} and a recent systematic review recommended against their use {9}. Further, respiratory tract infections can be more severe if resulting from aspiration of thickened liquids. Potential adverse effects and unintended consequences also include dehydration, urinary tract infections, thirst, oral and pharyngeal residue and impaired medication bioavailability {10}. Other issues include a lack of quality research evidence supporting the intervention and dislike of thickened liquids by people with swallowing problems, mainly because it impacts both their quality of life and enjoyment of drinking {11}. Informed decision-making deficits are also noted which impact treatment adherence {12,13}. Noting these concerns, the Royal College of Speech & Language Therapists (RCSLT) issued a position statement {10} and a subsequent paper {14} on the intervention of thickened liquids. The organisation advised against thickeners being used “as a blanket approach or go-to treatment” and highlighted the need for decisions to be made ‘through a process of informed consent following a holistic assessment that includes consideration of the potential impact on health and quality of life’ (p4).

In the annual year 2023/24, thickeners were the 7^th^ highest spend category out of all (35) nutrition borderline substance categories in the UK {15}. The US market in thickeners was worth circa $584.6 million in 2023 and is projected to rise to $874.1 million in 2033 {16}. Little data is available on prescribing or dispensing practices to support suggestions of decreased use, inform discussions and guide health professionals and policy makers in their practices and policy developments. It is also not known at present, whether the debate about appropriate use of thickeners has impacted prescribing/dispensing practices. In this study, we accessed the NHS Business Services Authority ePACT2 database {17} to examine dispensing trends over the last five years. While the information available to us regarding dispensing practices is currently limited to NHS England GP dispensing, this data will provide the first overview of thickener dispensing practices, serve as foundational material for parties interested in research, practice and policy making, and guide further work in this area.

### Aims

To explore the data on English GP dispensing of thickeners for the period 2019-2024 and identify dispensing practices and trends.

## Method

### Data collection

Every NHS prescription issued and dispensed for a general practice patient is entered onto the administrative database of the NHS Business Service Authority (NHS BSA), the ePACT2 (electronic Prescribing Analysis and Cost) {17}. The ePACT2 database is publicly available and comprehensive in managing NHS primary care prescribing and dispensing costs across England. Dispensed items are those items for which prescriptions were written by general practitioners and subsequently filled. Data was obtained to conduct an analysis of thickener dispensing on ePACT2 data with retrospective analysis performed on the anonymised data for the five-year period 2019-2024. Although ethical approval for use of this dataset is not required, the authors sought and received permission to publish this data in this paper from the NHS BSA after sharing analyses and results with this body. The data was downloaded onto an excel spreadsheet for analysis.

### Data analysis

Data were analysed descriptively. The primary outcomes of interest for this report were the linear trends in total number of thickeners dispensed in the different age groups. These was analysed using a linear regression calculator (Statistics Kingdom) {18}. R-squared - the percentage of the variance explain by the regression - was used to measure the goodness-of-fit of the model. The F test was used to check if the entire regression model is statistically significant.

## Results

Table 1 shows the breakdown of number of items dispensed, and the associated costs, by age-group. Between 2019 and 2024, nearly three million thickening items were dispensed in primary care at a cost of nearly 90 million sterling.

**Table 1:**
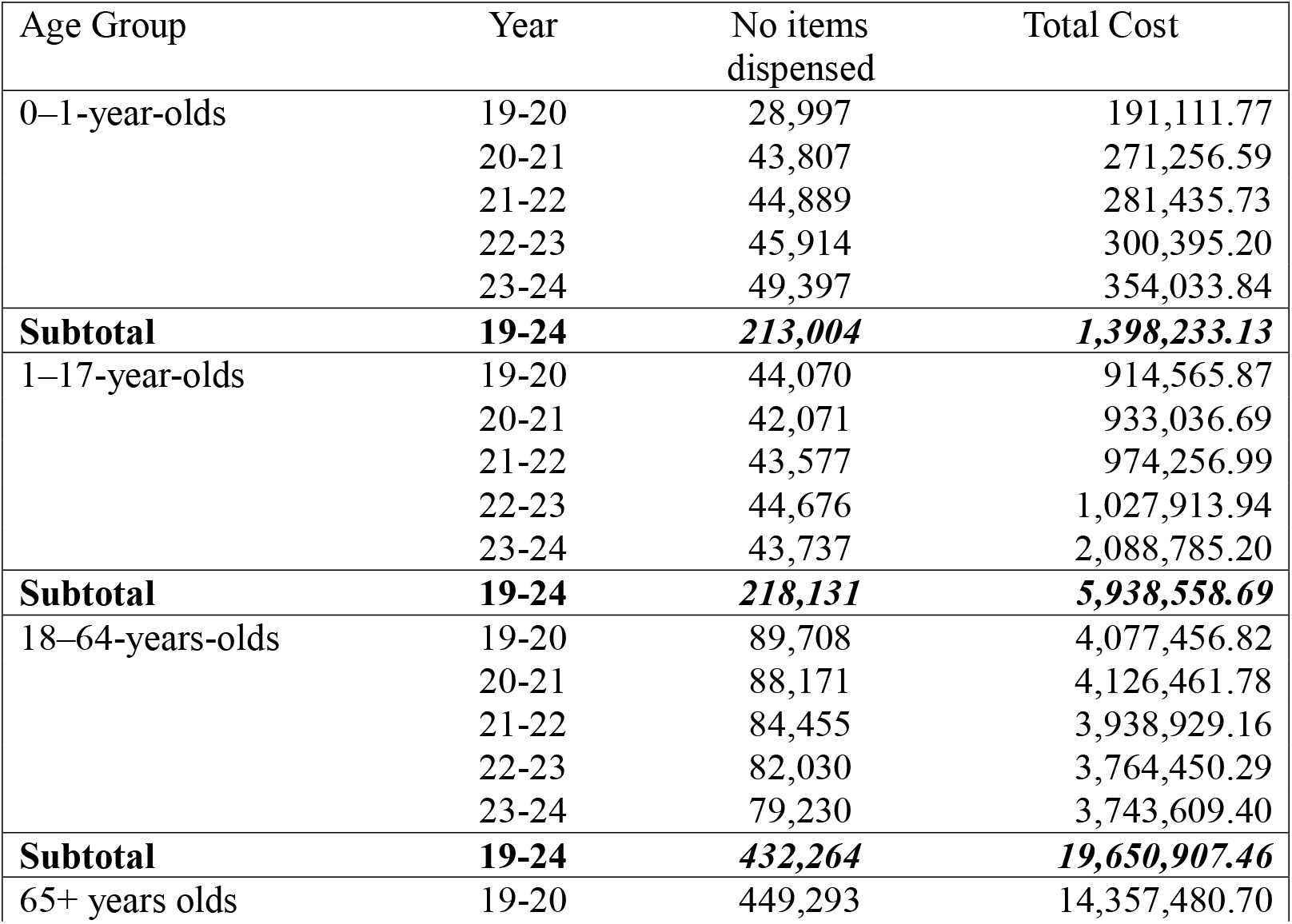
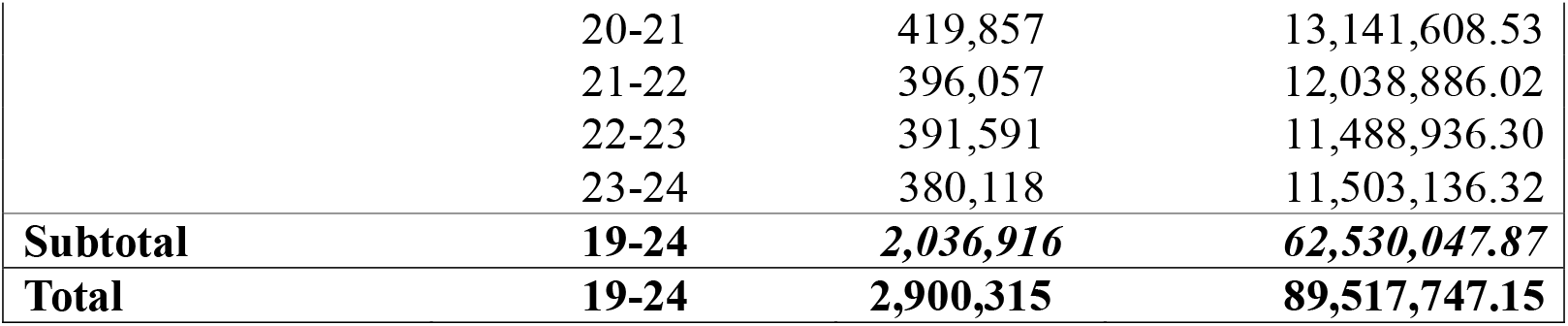
No. of thickening items dispensed based on GP prescriptions and age in England 2019-24

Figure 1 shows the total number of thickening items dispensed across all age groups. Approximately 612,000 items were dispensed in 2019-2020 and 552,482 were dispensed in 2023-2024, a 15% drop in dispensed items in the five-year period. Figure 1 shows a reduction in the total number of dispensed items year on year. Trends in dispensed items between 2019-20 and 2023-24 were different in the different age groups (Table 1).

**Figure 1.**
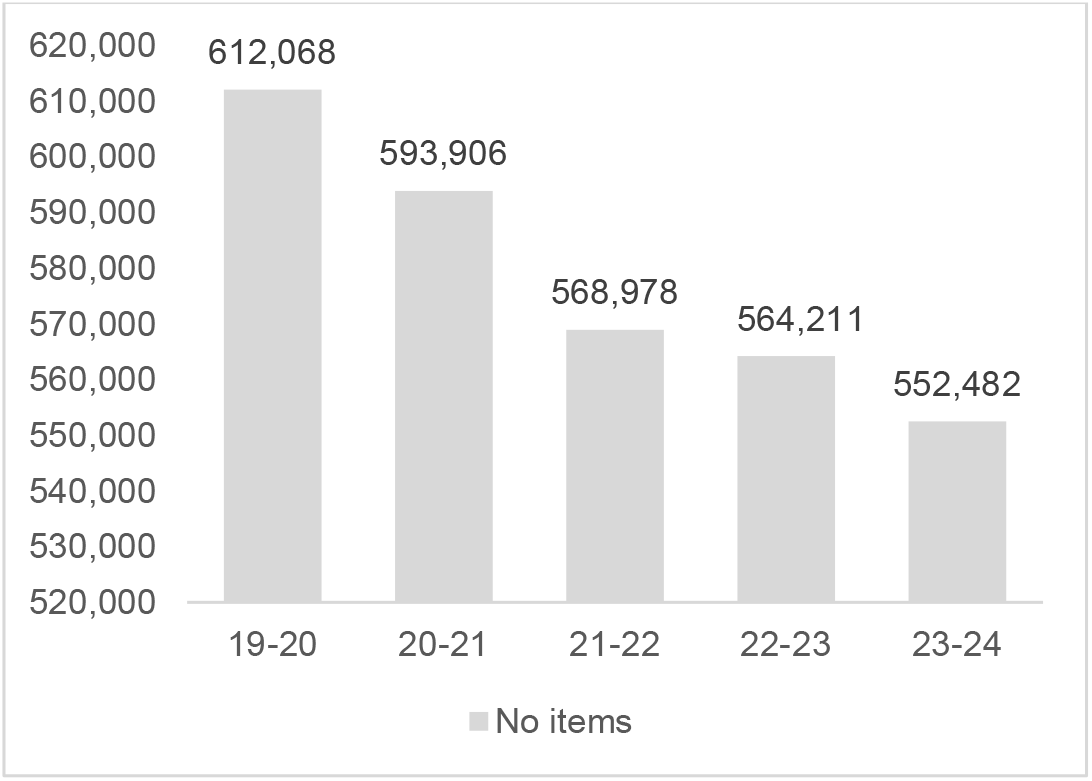
Total no of thickening items dispensed 2019-24

In the 0-to-1-year age group, which accounts for least dispensed items, there was a 70.4% increase overall in these 5 years. The linear regression equation for X (year) predicting Y (number of items) was Y = −8627490 + 4290X. R square was 0.74 (an R square of greater than 0.9 is regarded as high and ‘good’) showing that 74% of the variation in dispensed items was explained by the year. Overall, the linear model showed a non-significant increasing trend: F(1,3) = 8.52, p =.062.

In the 1-to-17-year age group, there was a negligible 0.7% increase in thickened items dispensed between 2019-20 and 2023-24. The linear regression equation was Y = - 376553.7 + 207.9X. R square was 0.12. There was no significant linear trend: F(1,3) = 0.4, p =.57.

In the 18-to-64-year age group, there was an 11.7% decrease in thickened items dispensed between 2019-20 and 2023-24. The linear regression equation was Y = 5561022.5 - 2709.7X. R square was 0.99, showing that the year explained almost all variation in dispensed items. The declining linear trend over 5 years was highly significant: F(1,3) = 271.53, p <.001.

In those aged 65 years or more, there was a 15.4% decrease in thickened items dispensed between 2019-20 and 2023-24. The linear regression equation was Y = 457368 - 16661.6X. R square was 0.92. This declining linear trend over 5 years was again significant F(1,3) = 32.41 (p = 0.011).

Figure 2 highlights that the older age group (65+ years old) accounts for most dispensed items, with more than two million thickening items (70.2%) being dispensed in primary care for older people at a cost of more than 62 million sterling.

**Figure 2.**
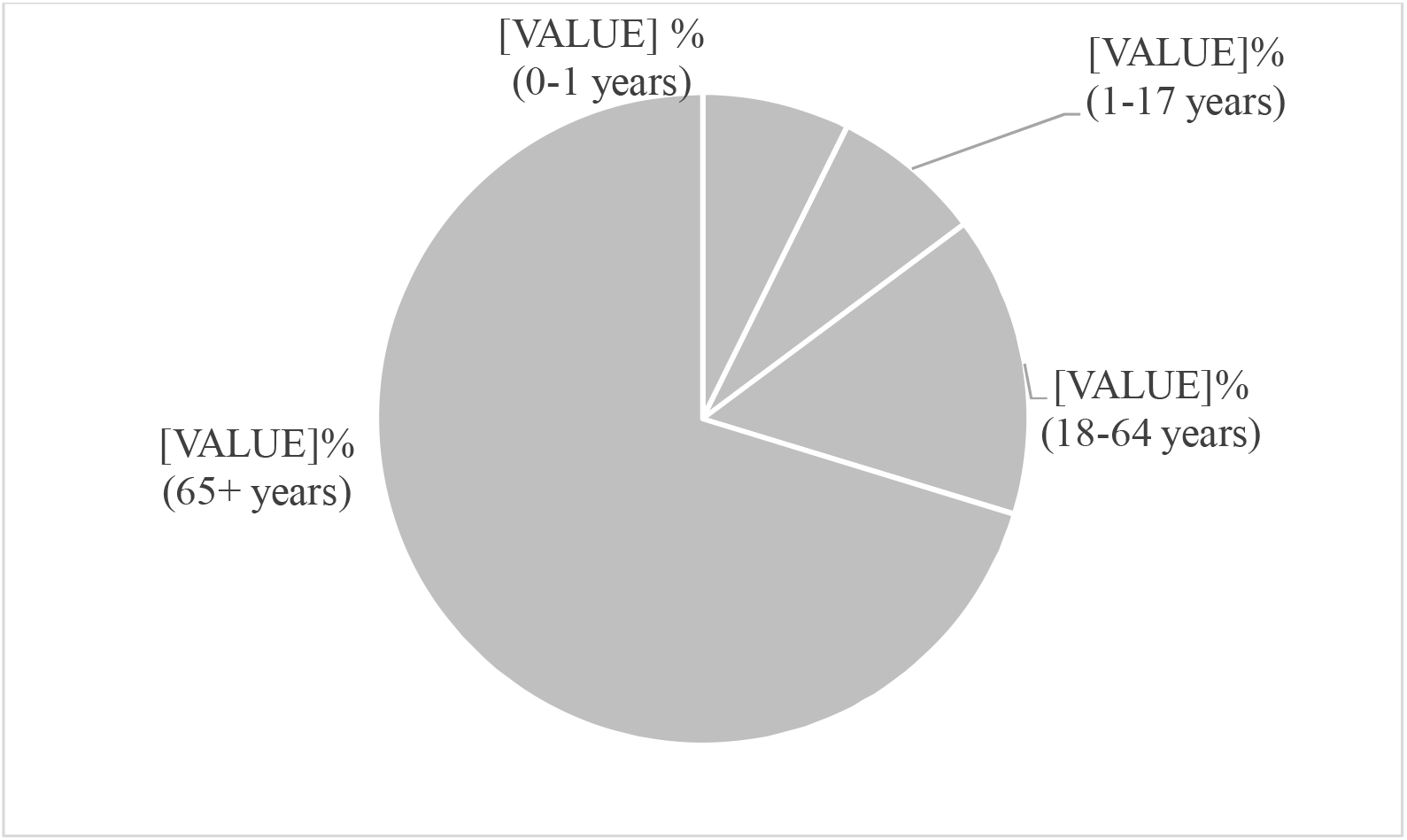
Percentage of total thickening items dispensed by age between 2019-2024

Figure 3 highlights cost of dispensed items by year. Despite a reduction in the total number of dispensed items and a decreasing trend from 2019-2-23, costs are seen to rise in the most recent year for which data is available. Although nearly 12,000 less items were dispensed in the 2023–2024 period across all age groups, constituting a 15% drop, dispensed items cost the NHS nearly 7% more in the 2023-24 period than for the previous year.

**Figure 3.**
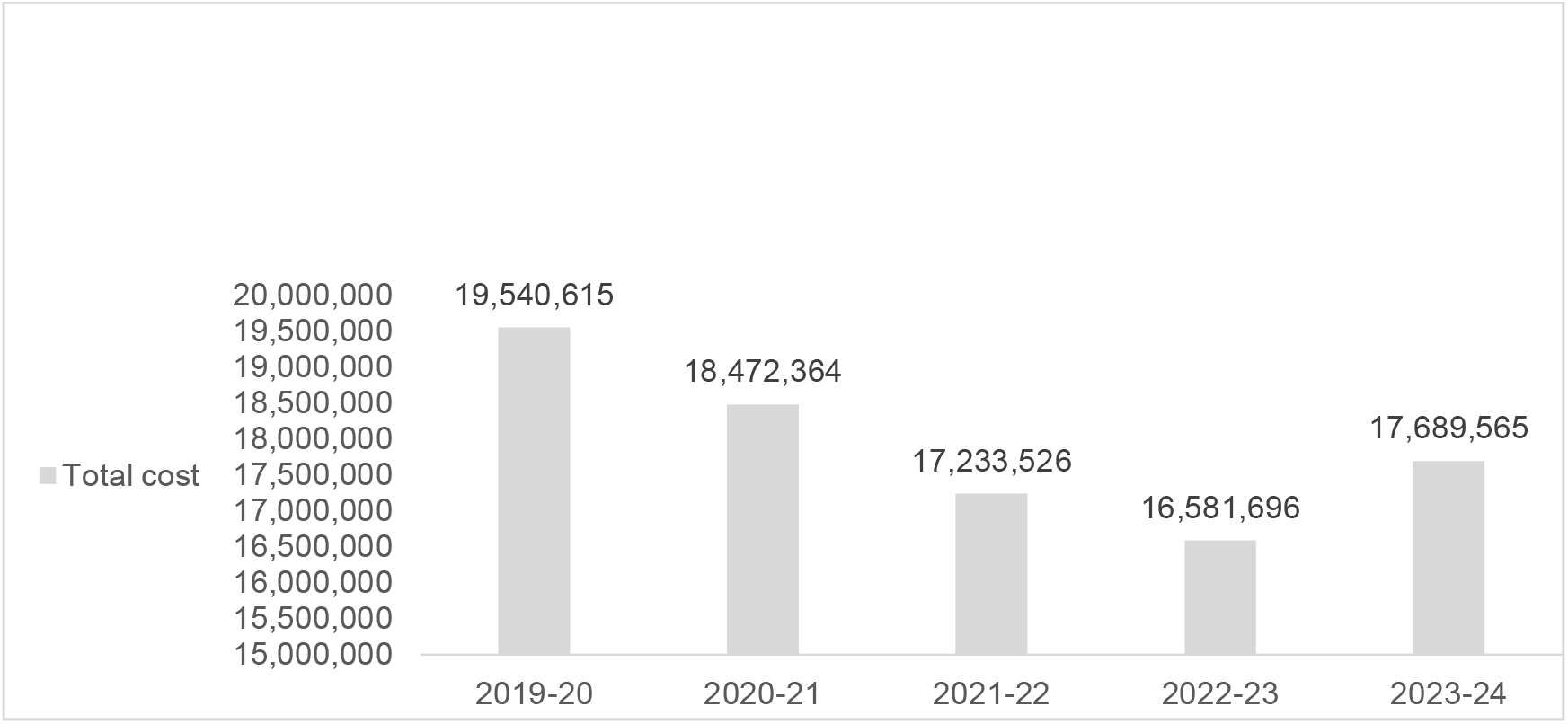
Total cost of thickening items dispensed 2019-24

Table 2 highlights price changes of the most dispensed items. Price increases vary between the selected products ranging from 6.6% to 36.8% (Table 2). Carobel products have shown the greatest price increase.

| Company/product | Price pre 2020 | Price as of 01/04/2024 | %increase |
| --- | --- | --- | --- |
| Thick & Easy | 5.41 | 6.10 | 12.75% |
| Resource Thicken Up | 8.46 | 9.02 | 6.6% |
| Nutlis | 5.50 | 6.75 | 22.7% |
| Nutlis Clear Sachets | 11.04 | 14.0 | 26.8% |
| Carobel | 2.91 | 3.98 | 36.8% |

Table 3 shows the number of costs of dispensed items by type. Of dispensed items over the five-year period, approximately 77% were gum-based, 12% were starch-based and 10% were carob-based. The cost of gum-based thickeners was higher than other products representing 87% of total costs (Table 3).

**Table 3:** Type of thickening items dispensed based on age 2019-24

| Age Group | Year | No. gum-based items | Gum-based costs | No. carob-based items | Carob-based costs | No. starch-based items | Starch-based costs |
| --- | --- | --- | --- | --- | --- | --- | --- |
| 0–1year-olds | 19-20 | 338 | 2,216.56 | 28,528 | 184,235.96 | 131 | 4,215.52 |
|  | 20-21 | 465 | 5,131.10 | 42,881 | 264,693.08 | 461 | 4,852.91 |
|  | 21-22 | 145 | 2,060.08 | 44,502 | 276,659.37 | 242 | 2,716.28 |
|  | 22-23 | 91 | 1,305.85 | 45,689 | 297,312.78 | 134 | 1,776.57 |
|  | 23-24 | 98 | 1,202.80 | 49,204 | 351,611.02 | 95 | 1,220.01 |
| **Subtotal** | **19-24** | **1,137** | **11,916.39** | **210,804** | **1,374,512.21** | **1,063** | **14,781.29** |
| **% of total** | **19-24** | **0.5** | **0.85** | **99.1** | **98.7** | **0.5** | **1.06** |
| 1–17year-olds | 19-20 | 16,946 | 525,823.09 | 16,981 | 169,630.30 | 10,143 | 219,112.48 |
|  | 20-21 | 17,405 | 571,341.27 | 16,910 | 177,878.88 | 7,756 | 183,816.54 |
|  | 21-22 | 19,523 | 627,810.44 | 17,032 | 176,477.55 | 7,022 | 169,969.00 |
|  | 22-23 | 21,201 | 688,927.53 | 17,699 | 196,594.51 | 5,776 | 142,391.90 |
|  | 23-24 | 22,518 | 777,858.86 | 17,481 | 213,274.61 | 3,738 | 1,097,651.73 |
| **Subtotal** | **19-24** | **97,593** | **3,191,761.19** | **86,103** | **933,855.85** | **34,435** | **1,812,941.65** |
| **% of total** | **19-24** | **44.7** | **53.7** | **39.5** | **15.7** | **15.8** | **30.5** |
| 18–64years-olds | 19-20 | 64,802 | 3,217,863.50 | 225 | 5,280.67 | 24,681 | 854,312.65 |
|  | 20-21 | 70,086 | 3,481,938.72 | 248 | 5,165.95 | 26,503 | 639,357.11 |
|  | 21-22 | 69,986 | 3,418,916.09 | 218 | 4,874.16 | 14,251 | 515,138.91 |
|  | 22-23 | 70,095 | 3,307,791.45 | 216 | 4,760.79 | 11,723 | 451,898.05 |
|  | 23-24 | 71,432 | 3,394,703.91 | 229 | 5,527.79 | 7,569 | 343,377.71 |
| **Subtotal** | **19-24** | **346,401** | **16,821,213.67** | **1,136** | **25,609.36** | **84,727** | **2,804,084.43** |
| **% of total** | **19-24** | **80.1** | **85.6** | **0.3** | **0.13** | **19.6** | **14.27** |
| 65+ years olds | 19-20 | 358,467 | 12,375,588.54 | 66 | 395.61 | 90,760 | 1,981,496.55 |
|  | 20-21 | 362,596 | 11,963,560.48 | 97 | 487.21 | 57,164 | 1,177,560.84 |
|  | 21-22 | 355,184 | 11,212,639.18 | 95 | 590.72 | 40,778 | 825,656.12 |
|  | 22-23 | 360,522 | 10,868,291.91 | 90 | 461.58 | 30,979 | 620,182.81 |
|  | 23-24 | 364,007 | 11,154,580.00 | 100 | 1,685.77 | 16,011 | 343,922.37 |
| **Subtotal** | **19-24** | **1,800,776** | **57,574,660.11** | **448** | **3,620.89** | **235,692** | **4,948,818.69** |
| **% of total** | **19-24** | **88.4** | **92.08** | **0.02** | **0.01** | **11.57** | **7.9** |
| **Total** | **19-24** | **2,245,907** | **77,599,551.36** | **298,491** | **2,337,598.31** | **355,917** | **9,580,626.06** |
| **% of total** | **19-24** | **77.44** | **86.7** | **10.3** | **2.6** | **12.3** | **10.7** |

Figure 4 shows the number of dispensed items by type across the five-year period under study. Carob-based dispensed items increased by approximately 68% over the period showing a large growth in dispensed items. This, combined with increased cost of Carobel products can be said to be a contributor to overall increased costs. Dispensing of gum-based items increased fractionally by nearly 3% while starch-based dispensed items decreased significantly by approximately 78%.

**Figure 4.**
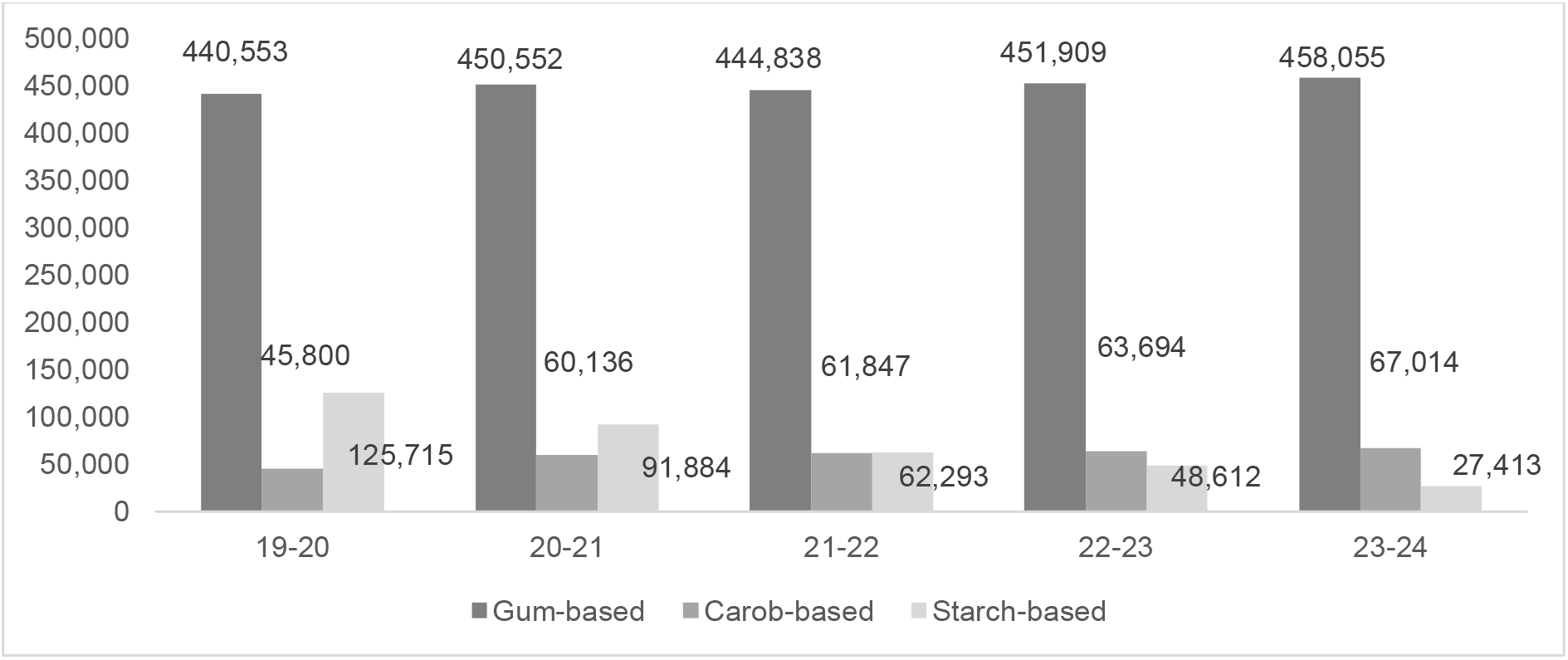
No. of thickening items dispensed by type 2019-24

Figure 5 shows the three main companies supplying dispensed items and the number of dispensed items. While a range of 33 items were dispensed from a range of companies during the last year for which data is available, Resource Thicken Up and Nutilis products, which represent the primary dispensed items in 2023-2024, account for 73% of dispensed products in that year.

**Figure 5.**
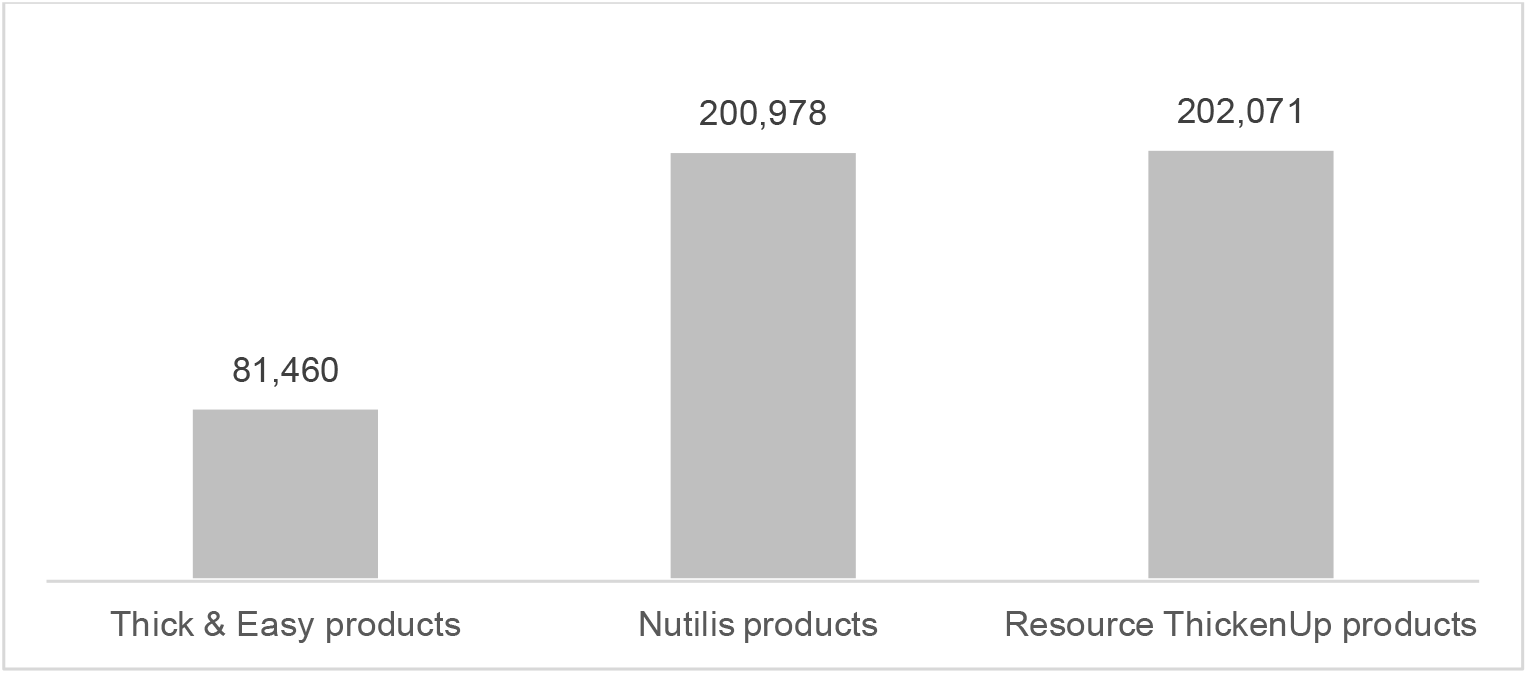
No of total items dispensed by primary suppliers 2023-24

## Discussion

Much has been written about thickeners over the last number of years, resulting in an RCSLT position paper on the topic {14} and the funding of a National Institutes of Health Research (NIHR) grant focused on de-implementation of the thickened liquid intervention. To date however, there has been a deficit of information regarding thickener prescribing/dispensing practices and this paper is intended to help rectify this situation.

### Number of items dispensed

Our results show a clear and significant decline in the number of thickening items dispensed in primary care between 2019 and 2024. While we only examined five years of data and this meant the a priori power of our study to detect a significant change was low, the decline was consistent and of such a magnitude, that our results are statistically significant. The data cannot explain why such a decline has occurred in England, however, the first of the ‘backlash’ papers on the topic in major journals were published from 2016 onwards {19-21} and has continued since. Awareness of the ongoing debate and (appropriately, in our view) a greater caution among SLTs in recommending and triggering prescriptions of thickeners seems a plausible explanation. Another possible reason for this decline may have its origins in SLT staff shortages with 19% of SLT posts in England vacant in the spring of 2024 {22}. The older population in England, the major recipient of dispensed thickeners, continues to rise rather than fall, and there has been no major decline in the prevalence of the common conditions that cause dysphagia. Hence these would not explain our findings. The years analysed included the years when the COVID pandemic was active and it is possible that there was reduced assessment and identification of dysphagia during this period, or that the pandemic led to the premature death of vulnerable older people with neurodegenerative conditions. However, this is unlikely to explain the ongoing decline in dispensing (and long COVID is a potential cause of dysphagia). The cost-of-living crisis may also be a contributory factor, especially given the NHS England decision to move many over the counter medicines off prescriptions. Most people under 60 now have to pay for prescriptions in England, and some may decide not to pick up some of their prescriptions because of cost {23}.

Of note, SLTs are not the only health profession recommending use of thickeners. There is limited evidence however, that nurse prescribers for example, are prescribing thickeners {4} and physician prescriptions continue to dominate prescribing in this area. Nevertheless, there is international evidence that thickeners are being used without the benefit of a clinical swallow exam being carried out by specialist dysphagia trained SLTs. This includes Ireland, where 70% of nurses reported that thickeners and modified texture diets were often started in the absence of a swallow exam performed by an SLT {24}; the USA where 80% of US SLTs encountered nursing-initiated texture modified practices {25}; New Zealand, where one third of residents (n=35,460) in long-term residential facilities were given texture modified diets without the benefit of an SLT consult or exam {26}, and; Canada, with health care assistants rarely accessing swallowing evaluations and demonstrating an overreliance on texture modified diets {27}. Estupinan Artiles et al.’s (2024) recent survey of Irish long-term residential settings highlighted that residents were provided with texture modifications including thickeners, if they had *suspected* swallowing problems {28}, while Bice et al.’s (2023) US-based study, provides evidence that two thirds of those receiving a modified solid/liquid diet in a continuing care setting (n=120), *did not actually have* an eating drinking and swallowing problem {29}. Thus, while a decline is noticed in GP dispensing, and the reduction in dispensed thickener items is a positive finding in our view, given the lack of evidence of benefit and the potential adverse effects, this trend may not reflect some practices in long-term residential settings, raising questions regarding overdiagnosis, overtreatment and treatment burden even in the absence of the condition for which the intervention is designed, and probably by individuals not trained in dysphagia. In the UK, food and drink thickeners are not tightly regulated as they are considered nutritional rather than medical products, which means they are subject to general food safety regulations rather than specific standards related to medicines.

### Age of people receiving dispensed items

The vast majority of community dispensed items - approximately 70% - were provided to older people and thickened liquids can legitimately be considered as an intervention primarily targeting older people. The data provided may be an underestimation given the LTRC practices cited above which may bypass SLT swallow exams and physician prescribing. Hammer-Castellanos et al. (2024) for example, in a survey of 253 US nursing homes, found a mean of 8.3% of residents were receiving the thickeners (with a high of 28% in some facilities) {30}. Irrespective, the high use of thickeners for this age group raises questions regarding the vulnerability of older people for this poorly evidenced, disliked and consequential intervention. Further, McGrail and Kelchner (2015) have shown that cognitive deficits predict oral fluid intake in people post-stroke receiving thickened liquids, raising valid questions regarding the use of thickeners in people with cognitive deficits {31}. There is good reason to believe that informed consent for use of thickeners is either not sought or may be based on a misrepresentation of the evidence. The dominance in thickener dispensing for the oldest age group is also interesting when considering epidemiological information. In England for example, 19% of the population is above 65+ years {32} and thus constitutes a small percentage of the population, but with a higher thickener dispensing rate.

Data on the youngest age group (0-1-years old) and in contrast to other age groups, highlights a significant rise in dispensed items over the five-year period, concurring with reports of witnessing a surge of thickener use with this population {33}. While the reason for this particular trend is unclear, in recent years, more SLT posts in neonatal facilities have been created which may have resulted in an increased focus on swallowing problems. More likely however, is that thickener use in this population is not limited to the treatment of swallowing problems but used for conditions such as gastro-oesophageal reflux disease (GORD) to reduce the number of regurgitation episodes {34}. This practice reflects guidelines which recommend thickening as the first-line approach to treat GORD in infants and young children {35}. Thus, the increasing trend of dispensed thickened items for the youngest age group, may have its roots not so much in swallowing problems, but in other medical conditions.

### Type of thickeners dispensed

The vast majority of thickeners dispensed were gum-based. Starch-based thickeners have significantly decreased as a proportion of dispensed items over the five-year period studied. According to the Specialist Pharmacy Service {36}, while starch-based thickening agents were widely used in the 1990s, gum-based thickening agents have gained popularity more recently. This is supported by the dispensing data. In contrast to starch-based thickeners, xanthan gum-based thickened fluids -in the UK, gum-based thickening agents are mostly xanthan gum-based-are reported to be effective in reducing aspiration for several reasons, including not increasing pharyngeal residue, better texture perception and better resistance to salivary amylase {37}. Calmarza-Chueca et al. (2022) also report better stability across time and temperature (6.5% vs. 43%) {38}, although Kim and Yoo (2018) have shown gum-based beverages thickening after a 15–45-minute prep window {39}. This is important as it is not uncommon, particularly with elderly people, for a thickened drink to be sitting for several hours. Further, Kim et al. (2017) showed that gum-based thickeners significantly altered sensory attributes {40}. Thus, questions remain on the quality and performance of gum-based thickeners despite their dominance of the market.

Carob-based dispensed items showed a large relative growth during this five-year period and carob seed/gum is used primarily for the youngest age group. It is reported to be a high viscosity product which has the property of absorbing water up to 40%, making it an effective thickener, stabilizer and emulsifier {41}. Carobel is the only ACBS approved thickener in children under 3 years old, with limited options being available due to the general high sodium content in thickeners. In this population, it is licensed for reflux and regurgitation rather than dysphagia. Once children reach the age of 3 years, it is recommended that they switch to gum-based thickeners {42}.

## Conclusions

While the data provided in this study is limited to GP dispensed thickeners in England, it provides researchers, clinicians and policy makers with initial information on dispensing practices. From the data it is evident that there is a significant decreasing trend in dispensed thickening items in adult populations and that the use of thickeners is primarily an intervention for older people. Thickener dispensing has increased in the youngest age group (0-1-years-old). The cost of thickeners to the NHS has risen in the last year in particular despite the significant decline in use observed for most age groups. From the available data, thickeners can be said to be primarily an intervention for older people and gum-based products dominate the market. This study raises a number of questions regarding the use of thickeners for people with swallowing problems.

## Limitations

The nature of the database used in the study means our data are highly reliable. However, there are limitations in the data also. It represents dispensed products only; more prescriptions may have been written but not dispensed, for example if the person chose not to take thickener. We cannot know whether dispensing was occurring in residential care or in the community or whether they were initiated by SLT recommendations. We do not know if differences exist based on geography. It would also be helpful to have data from earlier years to better judge whether the trajectory of decline can be attributed to the debate about thickened liquids. Our data are also too early to show any effect from the RCSLT publications, and it will be interesting to see whether the decline continues.

## Recommendations

1. A number of findings require further exploration, including the dominant use of thickeners for older people and the increase in use of thickeners for the youngest group, both of whom are particularly vulnerable groups.
2. It would be helpful to explore data from earlier years not included in this study, to better judge whether the trajectory of decline can be attributed to the debate about thickeners. It will also be helpful to repeat the review of the data in the coming years to identify any effect from newer RCSLT policy, and it will be interesting to see whether the decline continues.
3. The development of multiple accessible prescription/dispensing databases is required for improved knowledge and to support decision making and policy development. This includes, but is not limited to, geographical information, clinical conditions, prescriptions written vs. dispensed items, prescribing by non-GP physicians and sources of thickener recommendations. There is also a lack of data on how often prescriptions for thickeners are reviewed, and while the requirements are likely to change over time, this is not well documented and can result in under-ordered or stock piling and waste {43}.
4. Although there is an increased awareness among speech and language therapists regarding the debates surrounding thickeners, it seems likely that doctors and nurses will be less aware that the benefits of thickeners have been called into question. Education about thickeners targeting physicians, nurses, health-care assistants and care homes needs to be rolled out as a priority to establish an evidence-based and patient informed thickener culture, to improve safety and the care experience and to reduce overuse.
5. Continued research on gum-based thickeners is required given their dominance in dispensing practices including, but not limited to, physiochemical properties, safety and efficacy.

## Data Availability

www.nhsbsa.nhs.uk

## Disclaimer

The views expressed are those of the authors and not necessarily of the NHS BSA.

## References

1. Hong I, Bae S, Lee, H, et al. Prevalence of Dysphonia and Dysphagia Among Adults in the United States in 2012 and 2022. Amer J Speech-Lang Pathol. 2024 33(4), 1868–1879. 10.1044/2024_AJSLP-23-00407

2. Roberts H, Lambert K, Walton, K. The Prevalence of Dysphagia in Individuals Living in Residential Aged Care Facilities: A Systematic Review and Meta-Analysis. Healthcare. 2024 12, 649. 10.3390/healthcare12060649

3. Nielsen AH, Eskildsen SJ, Danielsen J, et al. Defining dysphagia – a modified multi-professional Danish Delphi study. Scan J Gastroenterol. 2023 58(6), 583–588. 10.1080/00365521.2022.2151850

4. Drennan VM, Grant RL, Harris R. Trends over time in prescribing by English primary care nurses: a secondary analysis of a national prescription database. BMC Health Serv Res. 2014 14, 54. 10.1186/1472-6963-14-54

5. Jones O, Cartwright J, Whitworth A, et al. Dysphagia therapy post stroke: An exploration of the practices and clinical decision-making of speech-language pathologists in Australia. Int J Speech-Lang Pathol. 2018 20(2), 226–237. 10.1080/17549507.2016.1265588

6. McCurtin A, Healy C. Why do clinicians choose the therapies and techniques they do? Exploring clinical decision-making via treatment selections in dysphagia practice. Int J Speech-Lang Pathol. 2017 19(1), 69–76. 10.3109/17549507.2016.1159333

7. McCurtin A, Byrne H, Collins L, et al. Alterations and Preservations: Practices and Perspectives of Speech-Language Pathologists Regarding the Intervention of Thickened Liquids for Swallowing Problems. Amer J Speech-Lang Pathol. 2024 33(1), 117–134. 10.1044/2023_AJSLP-23-00226

8. Painter V, Le Couteur DG, Waite LM. Texture-modified food and fluids in dementia and residential aged care facilities. Clin Int Aging. 2017 12, 1193–1203. 10.2147/cia.s140581

9. Hansen T, Beck AM, Kjaersgaard A, et al. Second update of a systematic review and evidence-based recommendations on texture modified foods and thickened liquids for adults (above 17 years) with oropharyngeal dysphagia. Clin Nutr ESPEN. 2022 49, 551–555. 10.1016/j.clnesp.2022.03.039

10. Position statement on the use of thickened fluids in the management of people with swallowing difficulties [Internet]. UK: Royal College of Speech and Language Therapists. 2023 [accessed 2025 Mar 4]. Available from: https://www.rcslt.org/wp-content/uploads/2023/03/Position-statement-thickened-fluids-1.pdf.

11. Swan K, Speyer R, Heijnen BJ, et al. Living with oropharyngeal dysphagia: Effects of bolus modification on health-related quality of life—A systematic review. Qual Life Res, 2015 24(10), 2447–2456. 10.1007/s11136-015-0990-y

12. O’Keeffe ST, Leslie P, Lazenby-Paterson T, et al. Informed or misinformed consent and use of modified texture diets in dysphagia. BMC Med Ethics. 2023 24(1), 7. 10.1186/s12910-023-00885-1

13. McCurtin A, Healy C, Kelly L, et al. Plugging the patient evidence gap: What patients with swallowing disorders post-stroke say about thickened liquids. Int J LangCommun Dis. 2018 53(1), 30–39. 10.1111/1460-6984.12324

14. Position paper on thickened fluids. [Internet]. UK: Royal College of Speech and Language Therapists. 2024 [accessed 2025 Mar 4]. Available from: Thickened fluids | RCSLT

15. NHS Business Services Authority [Internet]. UK. [accessed 2024 Sept 5]. Available from: www.nhsbsa.nhs.uk.

16. Dysphagia Diet Thickening Agents Market Outlook 2023 to 2033. [Internet]. USA. Persistence Market Research. [accessed 2025 Jan 20] Available from: https://www.persistencemarketresearch.com/market-research/dysphagia-diet-thickening-agents-market.asp

17. NHS Business Services Authority ePACT2 database. [Internet]. UK. [accessed 2024 Sept 5]. Available from: https://www.persistencemarketresearch.com/market-research/dysphagia-diet-thickening-agents-market.asp

18. Statistics Kingdom. [Internet]. [accessed 2024 Nov 8]. Available from: https://www.statskingdom.com/linear-regression-calculator.html.

19. Beck AM, Kjaersgaard A, Hansen T, et al. Systematic review and evidence based recommendations on texture modified foods and thickened liquids for adults (above 17 years) with oropharyngeal dysphagia - An updated clinical guideline. Clin Nutr. 2018 37(6A),1980–1991. doi:10.1016/j.clnu.2017.09.002

20. O’Keeffe ST. Use of modified diets to prevent aspiration in oropharyngeal dysphagia: is current practice justified? BMC Geriatrics. 2018 18(1),167. Doi.org//10.1186/s12877-018-0839-7

21. Wang CH, Charlton B, Kohlwes J. The horrible taste of nectar and honey— inappropriate use of thickened liquids in dementia: a teachable moment. JAMA Intern Med, 2024 176(6),735–6. 10.1001/jamainternmed.2016.1384

22. Vacancy survey. [Internet]. UK. [accessed 2025 Jan 17]. Royal College of Speech and Language Therapists, 2024, Available from: https://www.rcslt.org/policy-and-influencing/uk-wide/vacancy-survey/

23. Continuing to pay the price: The impact of prescription charges on people living with long term conditions. [Internet]. UK. Prescription Charges Coalition. 2023 [accessed 2025 Jan 21]. Available from: Prescription Charges Coalition - Home.

24. Oken M, Yen Chen K, O’Keeffe ST. “In Limbo”—use of, and alterations to, modified diets by nursing home staff in the absence of timely specialist support. 2024. Frontiers Rehabil Sci, 5. 10.3389/fresc.2024.1276713

25. Gurevich N, Osmelak DR. Speech-language pathologists’ experience with nursing initiated texture modified diets in health care settings. Adv Commun Swallow. 2024 27, 107–113. DOI:10.3233/ACS-240002

26. Miles A, Liang V, Sekula J, et al. Texture-modified diets in aged care facilities: Nutrition, swallow safety and mealtime experience. Austr J Ageing, 2020 39, 31–39. 10.1111/ajag.12640

27. Affoo R, Cliffe Polacco R, Lam B, et al. Dysphagia and Oral Health Concerns in Long-Term Care. Can J Speech-Lang Pathol Audiol. 2023 47(2), 109–124. CJSLPA_Vol_47_No_2_2023_1289.pdf

28. Estupiñán Artiles C, Donnellan C, Regan J, et al. Dysphagia Screening in Residential Long-Term Care Settings in the Republic of Ireland: A Cross-Sectional Survey. Dysphagia. 2024. doi:10.1007/s00455-024-10762-7

29. Bice EM, Galek KE, Ward M. Dysphagia and diets in skilled nursing facilities when patient’s health status changes: The role of imaging. J Amer Med Dir Assoc. 2023 25(2), 381–386. 10.1016/j.jamda.2023.11.008

30. Hammer Castellanos V, Butler E, Gluch L, et al. Use of thickened liquids in skilled nursing facilitiies J Amer Dietetic Assoc. 2024 104(8),1222-6, 10.1016/j.jada.2004.05.203

31. McGrail A, Kelchner L. Barriers to oral fluid intake: beyond thickened liquids. J Neuroscience Nurs. 2015 47(1), 58–63. doi: 10.1097/JNN.0000000000000114. PMID: 25565596

32. Population estimates for the UK, England, Wales, Scotland and Northern Ireland: mid 2022. [Internet]. UK. Office of National Statistics. [accessed 2025 Jan 17]. Available from: https://www.ons.gov.uk/peoplepopulationandcommunity/populationandmigration/populationestimates/bulletins/annualmidyearpopulationestimates/mid2022#:∼:text=1.-,Main%20points,from%2039.6%20years%20in%202011.

33. LaBarre Millet M. Overuse of Thickeners in the NICU. ASHA Leader. 2019 8–9. https://leader.pubs.asha.org140.234.253.9

34. Duncan DR, Larson K, Rosen RL. Clinical Aspects of Thickeners for Pediatric Gastroesophageal Reflux and Oropharyngeal Dysphagia. Curr Gastroenterol Rep, 2019 21(7), 30. doi:10.1007/s11894-019-0697-2

35. Rosen R, Vandenplas Y, Singendonk M, et al. Pediatric Gastroesophageal Reflux Clinical Practice Guidelines: Joint Recommendations of the North American Society for Pediatric Gastroenterology, Hepatology, and Nutrition and the European Society for Pediatric Gastroenterology, Hepatology, and Nutrition. J Ped Gastroenterol Nutr. 2018 66(3), 516–54. 10.1097/mpg.0000000000001889

36. Using thickeners of different types for patients with swallowing difficulties. [Internet]. UK. Specialist Pharmacy Service. [accessed 2025 Jan 17]. Available from:

37. https://www.sps.nhs.uk/articles/using-thickeners-of-different-types-for-patients-with-swallowing-difficulties/

38. Hadde EK, Mossel B, Chen J, et al. The safety and efficacy of xanthan gum-based thickeners and their effect in modifying bolus rheology in the therapeutic medical management of dysphagia. Food Hydrocolloids Health, 2021 1. 10.1016/j.fhfh.2021.100038

39. Calmarza-Chueca F, Sánchez-Gimeno AC, Raso-Pueyo J, et al. Rheological Properties and Stability of Thickeners for Clinical Use. Nutrients. 2022 14(17), 3455. doi: 10.3390/nu14173455

40. Kim CY, Yoo B. Rheological characterization of thickened protein-based beverages under different food thickeners and setting times. J Texture Studies. 2018 49(3), 293-299. Doi.org/10.1111/jtxs.12332

41. Kim H, Hwang H, Song K, et al. Sensory and rheological characteristics of thickened liquids differing concentrations of a xanthan gum-based thickener. J Texture Studies. 2017 48, 571–585. 10.1111/jtxs.12268

42. Cow & Gate Instant Carobel. [Internet]. Ireland. Nutricia Ireland. [accessed 2025 Jan 17]. Available from: Cow & Gate Instant Carobel.

43. Thickeners Guidance: NHS Norfolk and Waveney ICB Medicines Optimisation Dietetic Team (V3.1). [Internet]. UK. Norfolk and Waveney Integrated Care Board. 2024. [accessed 2025 Mar 3]. Available from: https://nwknowledgenow.nhs.uk/wp-content/uploads/2024/03/ThickenerGuidance_PPMO_Nutrition.pdf.

44. Trotman H, Ryeland S, Bradshaw S, et al. Buckinghamshire, Oxfordshire and Berkshire West Integrated Care System, Thickener Prescribing Guide for Primary Care and SALTs: Thickeners for patients with dysphagia. 2022. [accessed 2025 Mar 3].

45. https://www.bucksformulary.nhs.uk/docs/Guideline_901FMB.pdf

